# A flexible COVID-19 model to assess mitigation, “reopening”, virus mutation and other changes

**DOI:** 10.1101/2020.07.09.20150029

**Authors:** Sergio Bienstock

## Abstract

The COVID-19 epidemic which began in China last year has expanded worldwide. A flexible SEIRD epidemiological model with time-dependent parameters is applied to modeling the pandemic. The value of the effective reproduction ratio is varied to quantify the impact of quarantines and social distancing on the number of infections and deaths, on their daily changes. and on the maxima in these daily rates expected during the epidemic. The effect of changing R_eff_ is substantial. It ought to inform policy decisions around resource allocation, mitigation strategies and their duration, and economic tradeoffs. The model can also calculate the impact of changes in infectiousness or morbidity as the virus mutates, or the expected effects of a new therapy or vaccine assumed to arrive at a future date. The paper concludes with a discussion of a potential endemic end of COVID-19, which might involve times of about 100 years.

## Introduction

The epidemic of novel coronavirus disease (COVID-19) that began in China in late 2019 has expanded rapidly to over 220 countries and all U.S. states, and upended the lives and livelihoods of much of the world’s population. Over ten million cases and more than 500,000 deaths have been reported worldwide, of which 2.7 million and 128,000 respectively in the United States as of July 1, 2020 (1). The pandemic and associated community mitigation measures, have had a large negative economic impact as well. The virus could trim global economic growth by 3.0% to 6.0% in 2020, with a partial recovery expected in 2021 (2). Unemployment in the United States temporarily surged to levels not seen since the 1930s. Further, the epidemic is expected to exacerbate global poverty. It might push millions of people in the developing world, such as in India, Sub-Saharan Africa and South Asia, into extreme poverty (3).

Planning for the pandemic, keeping supply chains open, caring for the sick, and searching for effective therapies and vaccines have diverted the attention of healthcare professionals, researchers, planners, and essential workers around the world. Institutions such as the CDC in the United States are developing and refining COVID-19 scenarios designed to inform decisions by public health and other government officials (4). These aim at helping to evaluate the effects of mitigation strategies such as quarantines and social distancing, as well as with hospital resource allocation. Each scenario is based on a set of numerical values for the biological and epidemiological characteristics of COVID-19. These values — called *parameter values* — can be used in mathematical and statistical models to estimate the possible effects of the epidemic in different regions. They are being updated and augmented over time, as more is learned about the epidemiology of COVID-19.

Here I outline a differential equation-based SEIRD model with time-varying parameters and apply it to COVID-19 epidemic data. I discuss some of the potential implications of community mitigation strategies and their weakening in order to reopen the economy. This modeling approach is able to follow the entire development of an epidemic, from inception to eradication or endemic equilibrium, if desired. The model helps quantify tradeoffs in terms of additional cases and deaths in a given region and their maximum rates of increase, expected to result from a weakening of social distancing, or “reopening”. It can also be used to assess the impact of changes in a pathogen’s infectiousness or morbidity over time.

## Model and parameters

SIR and SEIR compartmental epidemiological models have been studied for nearly one hundred years. The term “compartmental” may have originated by analogy to trains or ships, essential modes of transportation in the first part of the twentieth century. In the non-autonomous SEIRD model discussed here, a population is divided into five non-overlapping classes, known as *compartments:*

- S, susceptible hosts;
- E, exposed hosts, presumed to be latently infected but not yet infectious;
- I, infectious hosts;
- R, hosts recovered from the exposed and infectious population and,
- D, hosts deceased due to the infection

This approach leads to a system of five ordinary differential equations in the compartmental variables, with time-dependent model parameters as coefficients. Shown in the Appendix, the equations describe the time evolution of each of the above-mentioned population groups. Key inputs to the model, defined in the Appendix, are the pathogen’s *basic reproduction number*, R_0_, or a closely related parameter, the *effective reproduction ratio*, R_eff_. From these the transmission rate from Susceptible to Exposed is calculated. Other required data are the transition rates from each group to the next. An E host can either get infectious (move to I) or recover (move to R), and an I host can either move to R (recover) or move to D (die from the disease). All Recovered hosts are assumed to be immune, or “removed” from the epidemic, although it is possible to relax this assumption. Hosts in all groups but D can also die for reasons unrelated to the infection, at an average rate. Finally, a gross birth rate for the Susceptible group is specified, and often set to the same value as the death rate unrelated to the infection (“steady state”). Such a SEIRD model would be described as an *open-population model in steady state*. All of the model parameters can vary with time, which affords the model a great deal of flexibility.

The starting point of the calculation (“time zero”) is specified by the fraction of the population, I, infectious at that time (typically very small, e.g. one per million) while S is nearly 1.0 for a new epidemic. Any other starting point is also possible. Expressed as fractions of the overall population, S + E + I + R + D should add up to 1.0 at all times for steady-state models. This provides a check on the accuracy of the numerical solution. All calculations were performed in the ***R*** Statistical Environment (5). The differential equations were easily integrated out to 100 years using the ode function in ***R*** (6) (7) (8) in order to explore the long-term behavior of the solutions. For practical uses, the focus is on the first year or two.

A rapidly growing set of reports on the COVID-19 infection parameters is becoming available online. Table 1 lists those used in this study (9) (10) (11) (12) (13) (14) (15) (16)

**Table 1.** COVID-19 model parameters.

| *Name* | *Symbol* | *Value* | *Comments* |
| --- | --- | --- | --- |
| Basic reproduction number | $R_0$ | 4.0 | The range 1-14 was examined. |
| Infection rate if exposed | $\sigma$ | 0.8/4.2 | Shown as probability/average period in days. |
| Recovery rate if exposed | $\varphi$ | 0.2/11.5 | Same as above. |
| Death rate if Infected (IFR) | $\omega$ | 0.0066/16.5 | Same as above. |
| Recovery rate if infected | $\Upsilon$ | 0.9934/11.5 | Same as above. |
| Death rate unrelated to COVID-19 | $\mu$ | 0.02/365 | Precise value immaterial in first years of infection. |
| Gross birth rate | $B$ | 0.02/365 | Same as above. |

The model allows the study of an epidemic from beginning to end. All of the model parameters can vary with time to reflect, for example, a change in social distancing, a mutation that alters the pathogen’s infectiousness, or a reduction in morbidity expected from a new therapy available as of some point in the future. Coupled with a modern differential equation solver (8), the model generates accurate results in seconds, which makes it feasible to perform sensitivity analyses with respect to one or more of the parameters used, as shown below in

Figure 3. Models based on local data could be run and the results easily aggregated as needed, in order to increase the granularity of predictions as data availability permits.

## Results

Figures 1 and 2 present the results of two runs with identical parameters, except as follows. In Figure 1 there is no attempt to slow down the infection rate with quarantine or social distancing, and R_eff_ is 4.0 throughout. The run in Figure 2 uses a R_eff_ of 4.0 up to 0.2 years (2.4 months) into the epidemic, at which point it goes down to 2.0 until 0.4 years (4.8 months), to simulate the potential effect of a quarantine extending for a bit over two months. A modified reopening with R_eff_ of 3.0 is assumed for all subsequent times. Shading is used to highlight the three time periods. The numbers inside each plot show the maximum value and/or end value of the dependent variable, as relevant. The last two plots in each figure are examples of *phase-space plots*, or *phase portraits* (17) (18); see also (19) and are discussed near the end of the paper.

**Figure 1.**
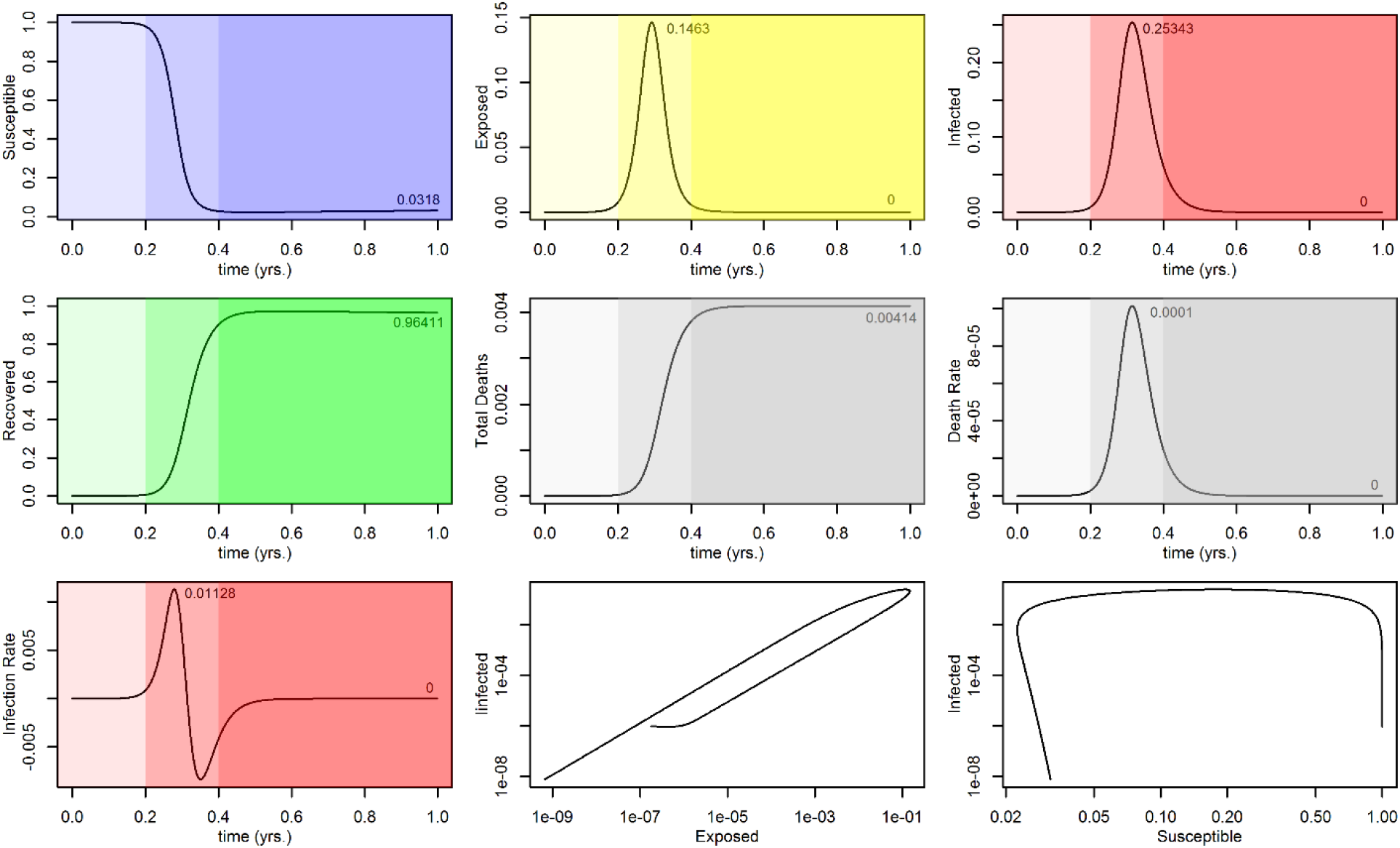
Label: No community mitigation assumed; R_eff_ =4.0 at all times.

**Figure 2.**
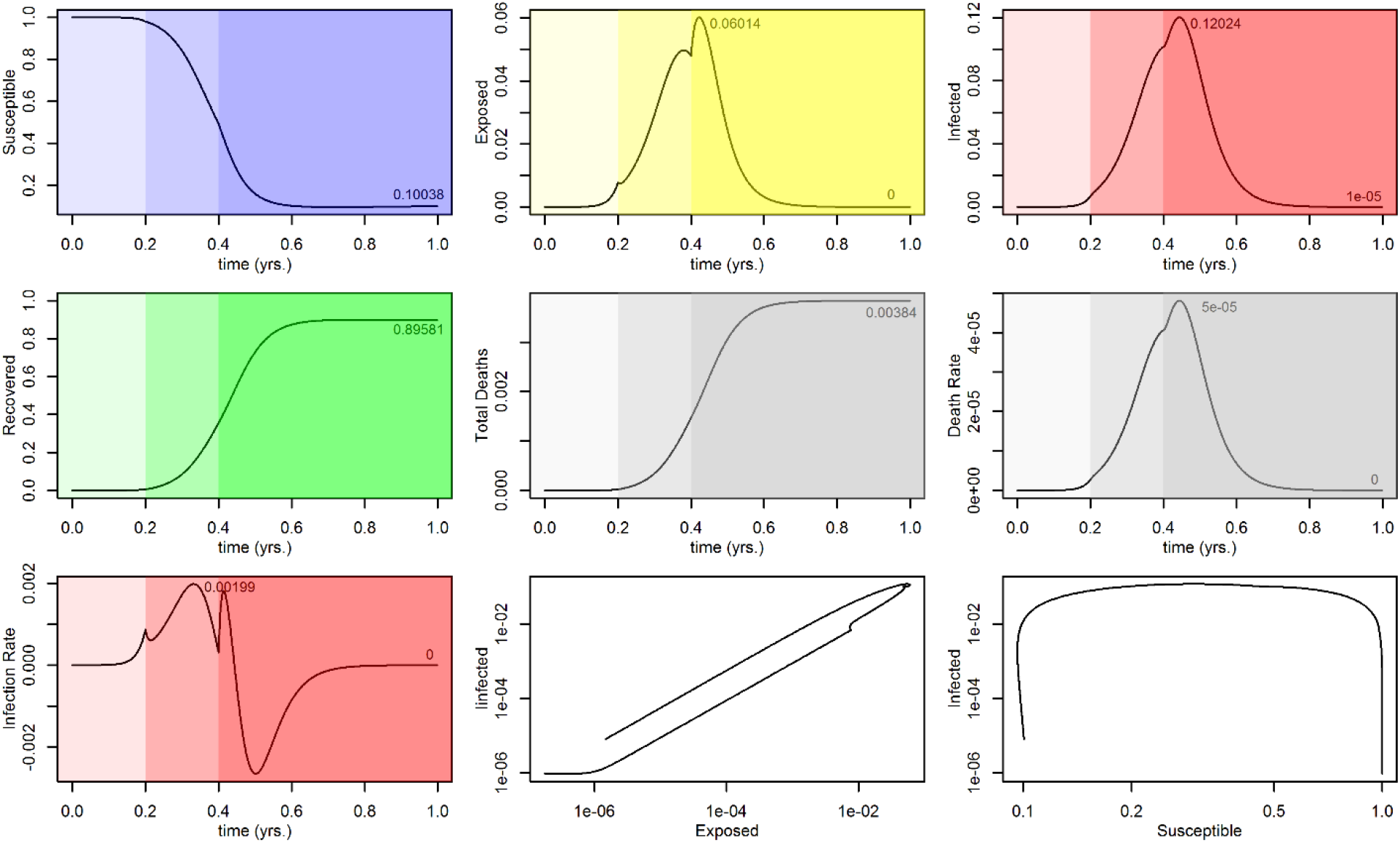
Label: R_eff_ values differ in the three shaded regions to simulate quarantine and reopening.

Each plot in Figure 2 should be compared with the corresponding plot in Figure 1. This shows that, applied at an opportune time, the quarantine succeeds in slowing down the infection (“flattening the curve”). The maximum values in the Exposed, Infected and daily death rate categories, all decrease by better than 50%. The impact on cumulative deaths and Recovered is less marked, as the epidemic is slowed down but not stopped (since R_eff_ remains greater than 1.)

Figure 3 shows the variation in the maximum daily infection rate with R_eff_ and with the time at which this maximum is estimated to occur. These are the curves in red. Because the infection probability increases with R_eff_, the larger maxima occur at correspondingly shorter times. The curves in blue show similar results for the estimated maximum daily death rate, or new deaths per day. As can be seen from Figure 3 and Table 2, the effect of a change in R_eff_ on these maximum daily rates is quite large. This type of information could be used to assess whether a relatively early weakening of quarantine and social distancing in a given region, leading to a substantial increase in R_eff_, would result in an acceptable number of additional casualties.

**Table 2.** Maximum daily infection and death rates vs R_eff_.

| $R_{\text{eff}}$ | $\max(dI/dt)$ | $1000 \max(dD/dt)$ |
| --- | --- | --- |
| 2.0 | 0.00189 | 0.03947 |
| 3.0 | 0.00634 | 0.07640 |
| 4.0 | 0.01128 | 0.10137 |
| 6.0 | 0.02050 | 0.13152 |
| 8.0 | 0.02835 | 0.14849 |
| 10.0 | 0.03504 | 0.15912 |

Figure 4 depicts the results of the run at R_eff_ = 4.0 when integrated out to 100 years. The use of logarithmic time as covariate allows the plot to cover a long time period and still show the early part of the epidemic in detail. Beginning at time near 30 years, *the model predicts a series of increasingly less severe resurgences of the epidemic*. Periodic and seasonal epidemic recurrences are well known and have been studied using SIR-class models, both autonomous and demographically forced (17) (18). The timing of the COVID-19 recurrences observed here varies with the birth and death rates used, *B* and µ. Why? Every year a certain proportion of the population dies for reasons unrelated to COVID-19, and a similar number of people are born, all of which into the Susceptible population — certainly true after maternal immunity, if any, disappears. Assume the disease is not eradicated, the pathogen has not lost its virulence, and no effective vaccine has been found. In time the Susceptible group becomes large enough for the virus to propagate again, albeit at a slower pace since some of the hosts it would encounter would be immune. Eventually the proportion of Susceptible and Infected hosts approach constant (equilibrium) values, at which point the epidemic becomes endemic. This is shown in the bottom right-hand plot of Figure 4, a phase portrait, where time increases as the curve is traced in a counterclockwise direction. Over the years the epidemic behaves like a damped oscillator, whose dampening is related to the variation in “herd immunity” over time. Endemic equilibrium is reached in approximately 100 years. Doubling *B* and µ would decrease the time to equilibrium by about half.

**Figure 3.**
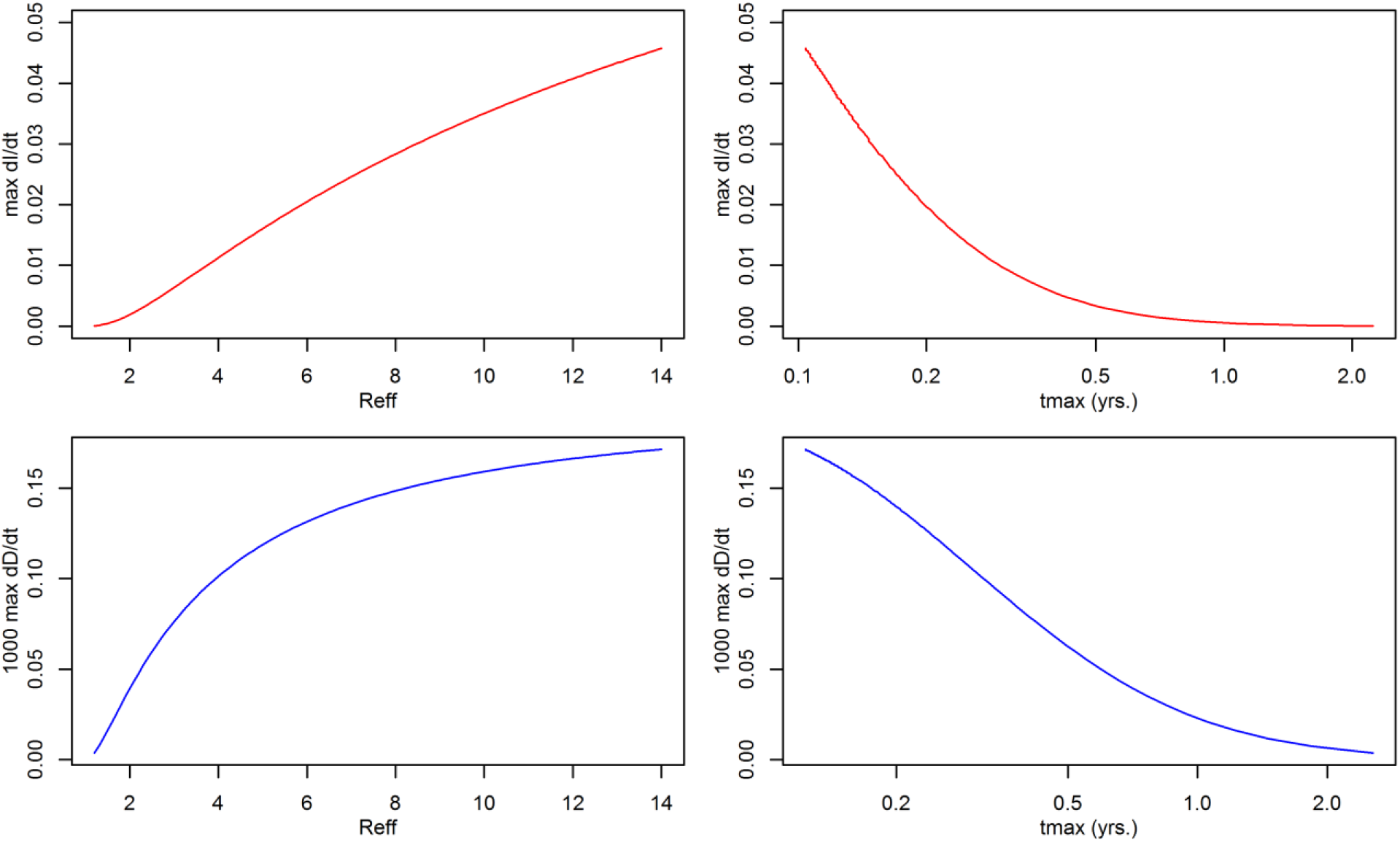
Label: Variation of maximum daily infection and death rates with R_eff_, and with time to maximum.

**Figure 4.**
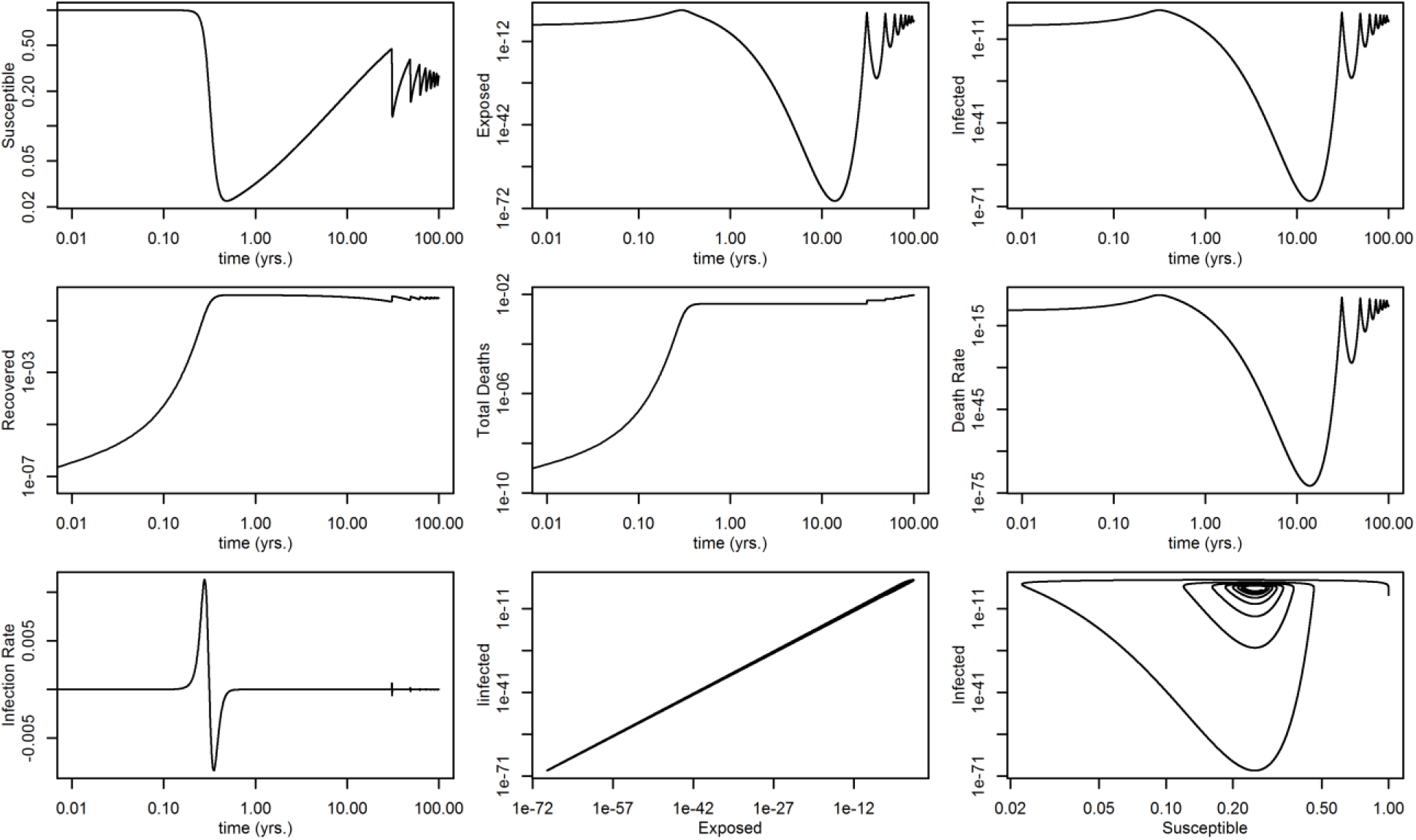
Label: Covid-19 epidemic modeled with R_eff_ =4.0 from inception to endemic equilibrium.

## Discussion

A SEIRD epidemiological model with time-dependent parameters was presented, able to follow an epidemic from inception to eradication or endemic equilibrium, and applied to COVID-19 data. By varying the value of the pathogen’s effective reproduction ratio, the model was used to assess the impact of quarantine and social distancing on the number of infections and deaths, on their daily changes (“new infections per day”, “new deaths per day”) and on the maxima in these daily rates expected during the epidemic. The effect of changing R_eff_ is substantial and ought to inform policy decisions around resource allocation to hospitals, appropriate mitigation strategies and their duration, and economic tradeoffs. Since all parameters can vary with time, the model is also able to quantify the effect of a change in the pathogen’s infectiousness or morbidity as the virus mutates, or the expected effects of a new therapy or vaccine arriving at some future date. Finally, the long-term potential endemic end of COVID-19 absent eradication is discussed which, the model suggests, might involve times of the order of 100 years.

## Data Availability

All data referred to in the manuscript has been published.

## Appendix SEIRD model diagram and its differential equations

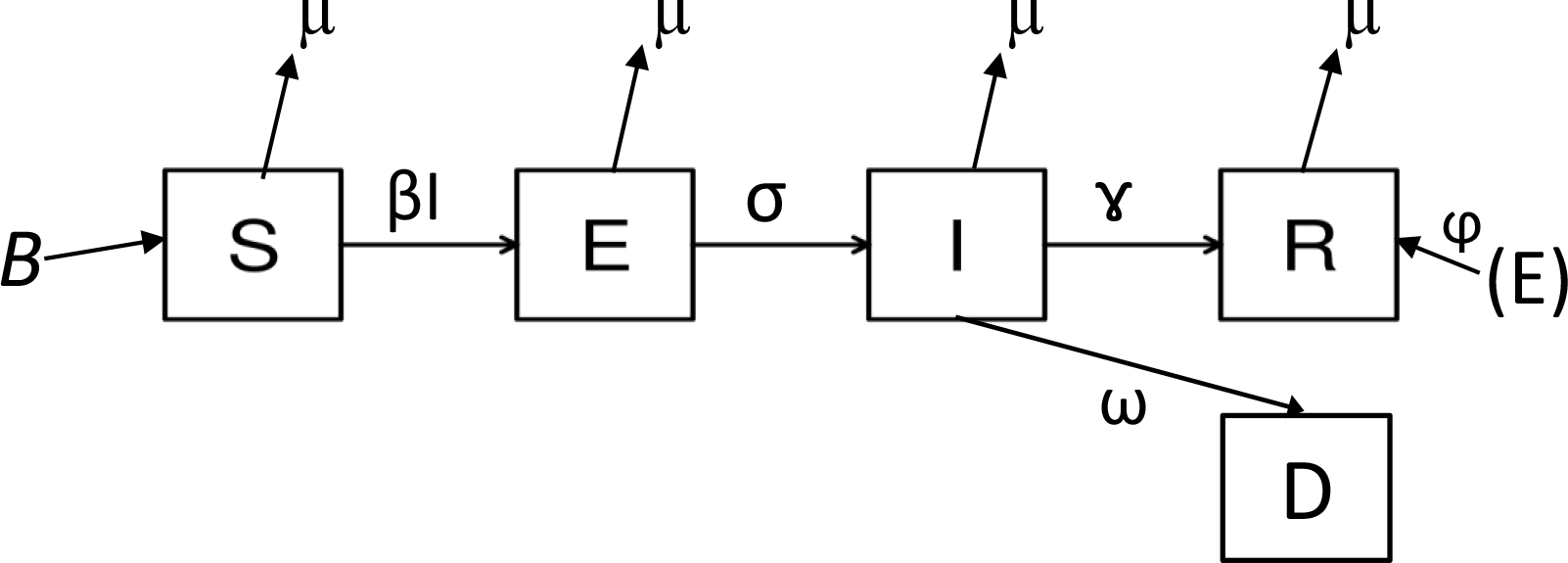

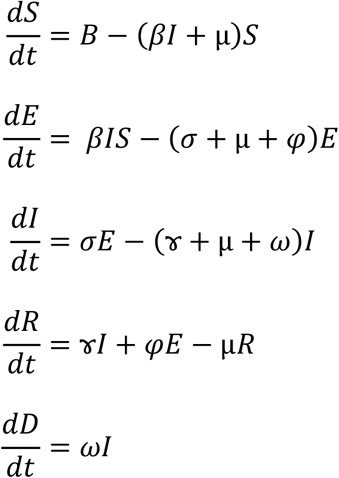

The basic reproduction number, R_0_, is the average or expected number of secondary cases one typical case would produce in a completely susceptible population (20) (21). The precise relationship between the transmission rate, β, and the pathogen’s R_0_ is model dependent. For the SEIRD model described, it is

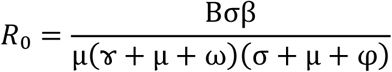

R_0_ has the following interpretation: it is the product of the production rates of E and I per unit contact, weighted by B**/**µ (22).

In this paper we define R_eff_ as R_eff_ = (1– *f*) R_0_ where *f* is the fraction of the Susceptible population that has been “removed” from the susceptible pool at a given time *though interventions* such as quarantine, social distancing, or immunity due to a vaccine. (The model already accounts for immunity in the Recovered group.) Other definitions of R_eff_ may differ. All of the model’s parameters can be arbitrary (known) functions of time.

## Notes

### Competing Interest Statement

The authors have declared no competing interest.

### Funding Statement

No funding supported this work.

